# Non-communicable disease care in peri-urban Nepal: Potential for community-based interventions

**DOI:** 10.1101/2025.08.01.25332626

**Authors:** Maryam Hameed Khan, Yoko Inagaki, Sonam Magar, Sweta Koirala, Nisha Rana, Pabitra Babu Soti, Gabriella Rose Chalker, Kamal Ghimire, Svea Closser, Dinesh Neupane

**Author notes:** Corresponding author: (DN). These authors contributed equally to this work (Maryam Hameed Khan, Yoko Inagaki).

## Abstract

Non-communicable disease (NCD) risk factors such as hypertension, diabetes, and smoking represent an increasing burden in Nepal, where access to equitable and affordable care remains limited. Task-sharing with Female Community Health Volunteers (FCHVs) offers a potential strategy to improve community-based NCD management, but its feasibility and scale up requires careful assessment. As part of the formative phase of the SCALE-NCD project, this qualitative study explored community and health system perspectives on NCD care and community-based delivery models in Pokhara, Nepal. Data were collected through 17 in-depth interviews and six focus group discussions with community members, FCHVs, facility-based community health workers (FB-CHWs), and organizational representatives. Thematic analysis revealed five core findings. Community members demonstrated awareness of NCD risk factors but reported deep mistrust in government health services, driven by negative experiences with public sector care, financial and structural barriers, and perceived discrimination. Perceptions of FCHVs were shaped by limited community exposure to their roles beyond maternal and child health. While some community members expressed skepticism about FCHVs’ capacity to manage NCDs, others valued their familiarity and accessibility, particularly when services were reliably supported. FCHVs and FB-CHWs emphasized that infrequent training, limited supervision, and chronic stockouts of supplies undermined service delivery. Financial strain and lack of consistent incentives also challenged FCHV retention and motivation. Stakeholders stressed that sustainable integration of community-based NCD care would require reliable government resourcing, local ownership, and alignment with existing systems. Participants also expressed cautious interest in mHealth strategies, such as SMS and audio messages, to support patient awareness and follow-up. These findings informed the final design of SCALE-NCD, a multi-component task-sharing intervention, and underscore the need to address structural constraints, institutional trust, and workforce support in scaling community-based NCD care programs in resource-limited settings.

## Introduction

Non-communicable diseases (NCDs), including hypertension, diabetes, and smoking-related illnesses, are a growing public health burden in low- and middle-income countries (LMICs), where health systems often struggle to provide effective long-term management. In Nepal, NCDs account for 71% of total deaths, with hypertension, high fasting blood glucose, and smoking among the leading risk factors for morbidity and mortality (1). Despite the rising burden, Nepal’s health system remains primarily oriented towards maternal and child health.

A key challenge in NCD care is the health workforce gap, particularly in LMICs where physician-to-patient ratios are low (2–4). Task-sharing approaches, where community-based providers take on expanded roles in prevention, screening, and referral, offer a promising strategy to bridge these gaps in NCD care (5–8). In Nepal, Female Community Health Volunteers (FCHVs)— a nationally deployed cadre of community health workers— represent a uniquely positioned workforce, embedded within their communities and experienced in health promotion. FCHVs are local women who serve on a voluntary basis and are not salaried by the government, although they may receive small allowances for specific activities (2). Previous research has demonstrated that FCHVs can effectively contribute to NCD care by measuring blood pressure, providing counseling, and referring high-risk individuals (2,9–11). However, despite growing interest in integrating FCHVs into NCD management, evidence on their role in long-term NCD care remains limited, particularly regarding expanding responsibilities, and the social and structural barriers to sustainable implementation (2,12,13). Equally important is understanding the broader structural and contextual factors, such as trust in government health systems, determinants of NCD risk prevention and management, and health equity considerations, that may shape the success of community-based NCD interventions.

In addition to task-sharing, while mobile health (mHealth) interventions, such as SMS reminders and phone call follow-ups, have demonstrated potential in supporting chronic disease management (14), their implementation in LMICs requires careful consideration of contextual factors. In Nepal, where digital literacy and phone ownership vary widely, early studies suggest potential for mHealth strategies, but evidence on their broader feasibility and acceptability in NCD management remains limited (15,16). To design effective and scalable interventions, it is critical to assess whether mHealth approaches can complement community-based health services.

This study presents findings from the formative, pre-implementation phase of the ‘Scaling Up Community-based Noncommunicable Disease Research into Practice in Pokhara Metropolitan City of Nepal’ (SCALE-NCD) project, a hybrid type 2 effectiveness-implementation study evaluating a task-sharing intervention with FCHVs for management of hypertension, type 2 diabetes, and tobacco smoking. Using qualitative data from diverse community and health system stakeholders, this paper explores the structural, social, and contextual factors that inform intervention design and delivery for community-based NCD care.

This study shows that systemic issues—such as trust in government health services, structural inequities in access to care, and sustainability challenges for volunteer-led models—may constrain the effectiveness of task-sharing approaches if left unaddressed. It also provides a grounded understanding of the opportunities and limitations of mHealth strategies in settings with variable digital literacy, and highlights both the promise and constraints of involving FCHVs in NCD care—particularly around sustained training and resources, adequate compensation, and stronger integration into the health system.

### Methodology

This qualitative study was conducted in Pokhara Metropolitan City, Nepal, in partnership with the Nepal Development Society (NeDS), a local community-based research and implementation organization. The study area comprises both peri-urban and rural communities with mixed reliance on public and private health systems.

Data collection took place in November 2024 through focus group discussions (FGDs) and in-depth interviews (IDIs) with key stakeholders, including:

1. Community members with NCD risk factors (i.e. hypertension, diabetes, or smoking) (five FGDs)
2. FCHVs (two IDIs, one FGD for a total of nine participants)
3. Facility-based community health workers (FB-CHWs) (six IDIs)
4. Representatives from key organizations, including government health agencies, local public health offices, academic institutions, international organizations, and telecommunications and advocacy sectors (nine IDIs)

Interviews were held at the NeDS office, health posts, or respondents’ homes in Nepali and lasted between 30 minutes and two hours. Written informed consent was obtained from all participants. Audio recordings were transcribed verbatim, translated into English, and supplemented by field notes to capture contextual details.

Thematic analysis was conducted using NVivo software. The analysis followed both an inductive and deductive approach. Deductive coding was informed by the study’s objectives to identify themes related to feasibility and implementation outcomes, and inductive coding allowed for the identification of emerging themes based on participant narratives.

The study received ethical approval from Johns Hopkins Bloomberg School of Public Health and the Nepal Health Research Council’s Institutional Review Boards.

## Results

### 1. Community composition

The study sites were located approximately 30–45 minutes from downtown Pokhara and encompassed a mix of hilly settlements and valley communities with varying degrees of urbanization. Neighborhoods featured a combination of newly built concrete homes, older clay or stone houses with tin roofs, roadside shops operating out of family homes, and pockets of fields dotted with haystacks, goats, and rice paddies. Some areas had densely built homes, while others—especially those uphill—were sparsely populated. In many neighborhoods, subsistence farming coexisted with the construction of modern infrastructure and increasing traffic. This geographic and demographic diversity shaped the mixed social composition of the communities described below.

The communities in the study area were diverse, with a mix of ethnic groups, socioeconomic classes, and occupational backgrounds. Some areas were historically home to Brahmin and Chhetri families with ancestral land and wealth, while others had a higher concentration of Dalit and Janajati groups.

While some respondents noted that sometimes there were economic disparities between caste groups, several participants emphasized that wealth was not solely determined by ethnicity. “*Some Dalits are wealthier than Brahmins in our area,”* an FCHV noted in a focus group. *“Some of these Dalits have significant land holdings and are doing well.”*

The community included people engaged in various professions, including farming, small business ownership, government jobs, and foreign employment. Respondents noted that older residents with inherited land and property tended to be better off, whereas newer migrants or those without ancestral holdings often faced economic hardships. Many younger individuals had migrated abroad for work, while older residents and women were more likely to be engaged in local occupations such as farming or running small businesses. Some families who had engaged in foreign employment or business had accumulated wealth, while others continued to struggle. In some areas, entire households had migrated, leaving homes empty or occupied only by elderly couples. Thus, there was a mix of areas that were urbanizing and areas that were emptying. *“If we go slightly uphill from here, many houses are completely empty*,” commented a health worker at a health post, gesturing towards the forested hillside. *“Just elderly couples. In some cases, even they have left.”*

### 2. Community perspectives on NCDs

#### Perception of NCD risk factors

Many participants attributed NCDs to unhealthy dietary habits and sedentary lifestyles. Processed and pesticide-laced foods were seen as unavoidable due to reliance on market produce, and excessive consumption of salt, oil, and meat was commonly cited as a risk factor. As one community member explained, *“For me, it’s because I wasn’t doing exercise. I ate unhealthy food, and in the evening, I’d sit around. Then, the next morning, I’d just sit at the shop.”* Health workers echoed this view, with one noting, *“Now, people have a sedentary lifestyle and don’t have daily work and exercise. So, the main reasons are dietary habits and exercise.”*

Community members suggested that wealthier individuals were more likely to develop NCDs due to their sedentary lifestyles, dietary choices, and lower engagement in physical labor. However, a few participants felt lower-income individuals were at risk due to limited awareness of preventive health measures and difficulty accessing health services.

Many participants also cited the role of alcohol consumption and smoking in the development of NCDs. A community member reflected, *“Alcohol, cigarettes, fatty and greasy foods. I ate too much of everything, more than I should have. That’s why I got it [hypertension]”.* While some participants briefly mentioned alcohol and smoking as contributing factors to NCDs, there was limited in-depth discussion on smoking-related behaviors or cessation.

Beyond lifestyle behaviors, many participants associated stress with the onset of hypertension, linking it to work pressure, family responsibilities, and financial stress. “*When you’re a parent, you have to look after the children. There’s a lot of worry involved—whether the child will go astray, whether they’ll earn or not, whether something will go wrong. Constantly worrying, those kinds of pressures start to build. Mentally it takes a toll,”* a participant noted.

A few community members also attributed their NCDs to previous physically demanding work experiences, particularly abroad. One individual recounted, *“For me, it was working in the Gulf countries… The heat from the fire would be intense. It’s like a pressure in that heat, and eating a lot of chicken and mutton, it added up. That’s why my pressure started to rise.”*

While lifestyle choices were frequently cited, a few participants also mentioned a genetic predisposition to NCDs*, “I think it’s hereditary. My parents had it, and now I do too.”* Genetics was particularly cited as a reason that, despite maintaining healthy behaviors, some individuals still developed hypertension and diabetes.

There were mixed opinions on whether men or women were more affected by NCDs. Some health workers stated that hypertension and diabetes were more common among men. However, another suggested that both genders were now at similar risk due to changing lifestyles. Some noted that while hypertension was more prevalent in men, women were more likely to visit health facilities for check-ups and follow-ups.

A few participants commented on differences in NCD risk among ethnic groups, particularly noting that certain groups had higher consumption of alcohol and tobacco, which might contribute to their risk. However, others stated that, in general, all ethnic groups were equally at risk for NCDs.

#### Barriers to management

##### I. Concerns about treatment

Concerns about hypertension and diabetes treatments led some individuals to delay or avoid medical intervention. Health workers said that a widespread belief in the community was that once someone starts taking hypertension medication, they must continue it for life, and that this discouraged individuals from initiating treatment, fearing long-term dependency.

Some community members expressed concerns about the medication. One commented, *“When you take medication for diabetes, it leads to other health problems… Diabetes medication causes other diseases. It’s very harmful.”*

##### II. Hesitancy to screening

FCHVs also encountered resistance from individuals who doubted the reliability of screenings. One FCHV commented, *“Some people said, ‘If you’re not going to give me medicine, why are you even testing me?’* Another commented, *“Some people didn’t cooperate. But when we found cases with higher risks and shared real-life examples, people became more conscious.”*

Several officials emphasized the high proportion of undiagnosed NCDs. A public health officer described findings from screening camps in Pokhara where 3-5% of attendees were newly diagnosed with hypertension or diabetes, and another confirmed the high burden of cases in communities. He noted “Whatever community we go to … a lot of hypertension patients can be found who are not taking medicine”, and another added that large portions of the population “remain hidden from the health systems and untreated.”

##### III. Delayed healthcare-seeking behavior

Even when diagnosed with hypertension or diabetes, some individuals remained skeptical of their test results, particularly due to the often-asymptomatic nature of chronic diseases. Health workers noted that some patients outright refused treatment because they did not feel unwell. One shared, *“Occasionally, when a patient’s BP is extremely high, like 200 or 210, and we provide treatment, they might say, ‘I don’t feel any symptoms. You’re saying my BP is this high, but I don’t believe it.’”*

Concerns about test accuracy further fueled mistrust. Some community members felt that readings taken at local medical stores were unreliable, leading them to question their diagnosis or delay follow-ups. *“They might say, ‘It’s 140/90,’ but when I go to a proper doctor or hospital later, it turns out to be 160/95.’”* a community member commented in an FGD.

Some community members reported checking their blood pressure periodically, typically every one to three months, either at a pharmacy, hospital, or health post. However, a few individuals mentioned that they only sought checkups when they experienced symptoms such as dizziness or discomfort, rather than proactively monitoring their condition at regular intervals. Some admitted they did not monitor their blood pressure or blood sugar regularly, relying instead on medication to keep their condition stable*, “There’s no fixed routine. If I feel unwell, I go to a medical store or hospital and get it checked”,* a participant explained in a community FGD.

Several FCHVs noted that reluctance to seek care often stemmed from individuals dismissing the severity of their condition. They recalled cases where neighbors ignored medical advice, and one shared, *“A neighbor, a few houses away from mine, came for a test. He had high sugar and blood pressure levels. When I measured his blood pressure, it was over 180. I told him to go to the doctor, but he ignored the advice. Within a week, he passed away.”* Another shared, *“I advised her to start treatment, but she brushed it off, saying, ‘I’m fine; nothing will happen to me.’ Within a year and a half, she too passed away.”*

##### IV. Financial barriers and access to care

The cost of treatment and follow-up care was a significant obstacle, particularly for lower-income individuals. Many FCHVs shared instances where people expressed frustration over the unaffordability of treatment. A health worker working in a health post recounted, *“One individual said, ‘These medicines are too expensive, I won’t buy them,’ and stopped taking diabetes medication.”*

Limited medication availability and frequent stockouts at government health facilities exacerbated these challenges, often forcing individuals to purchase medicines from private pharmacies at higher costs. Some people avoided check-ups altogether because of consultation fees and follow-up costs.

#### Treatment adherence

##### I. Lifestyle modifications

Upon diagnosis, some individuals reported making lifestyle changes to better manage their health. Respondents commented that they changed both exercise and dietary habits, such as reducing salt and sugar intake, exercising regularly, and avoiding alcohol. A community member shared, *“For diabetes, I don’t eat sugar at all. I have one meal of rice and another of roti. I avoid cold drinks. I used to drink alcohol, but I stopped after being diagnosed with diabetes.”*

However, long-term adherence to lifestyle modifications remained a challenge, with some reverting to previous habits over time, and a few commenting that they had not changed their diets despite being prescribed with medication. *“I used to eat the same things before I started taking medication. Now, I still eat the same way even with medication.”*

Other participants noted that although they attempted lifestyle changes initially, maintaining them consistently was difficult. Healthcare workers emphasized that while lifestyle modifications were frequently recommended to manage hypertension and diabetes, many individuals struggled to make these changes.

##### II. Medication adherence

Many participants stated that they had been taking their prescribed medication consistently for several years, highlighting a strong commitment to managing their conditions. Some individuals had been on medication for over a decade, with one participant noting that he had been taking hypertension and diabetes medication for 41 years without interruption.

While reports of continuous adherence were common, some individuals mentioned missing doses occasionally, either due to running out of medication or personal oversight. In some cases, individuals stopped taking their medications altogether, particularly if they felt better or assumed their condition had improved, or due to financial constraints. Health workers confirmed these views.

These challenges in managing NCDs were further compounded by widespread perceptions and experiences that shaped community trust—or mistrust—in the health system. While these findings highlight evolving risk awareness within the community, they also raise a critical question: where do individuals turn when they seek care?

### 3. Health system trust

#### Government health services

Trust in government health services was a recurring theme among participants, with many expressing skepticisms about the quality of care, availability of medications, and overall reliability of public healthcare facilities. While a few participants noted improvements in certain facilities, the general sentiment reflected a deep-rooted distrust in the public healthcare system.

##### I. Medication and supply shortages

One of the primary factors eroding trust was the frequent unavailability of essential medications, which were often out of stock or provided in insufficient dosages, forcing patients to purchase them from private pharmacies at higher costs. Even when medicines were available, supplies were often limited, requiring frequent return visits. As one participant commented, *“Sometimes they would give me the medicine, but other times they would say, ‘You have to buy it yourself.’”* Another added, *“At our local health post, they only prescribe a two-month supply of medicine. Once it’s over, we have to go back, pay, and get the prescription renewed.”*

Healthcare workers acknowledged these challenges, noting that frequent stockouts limited their ability to provide effective care. One health worker explained, *“At that time, there was a severe shortage of medicines. Even if we diagnosed with the issue [health condition], we didn’t have the capacity for first-line treatment at our level.”* They also noted that health posts provided only basic medications, with limited dosages that were insufficient for managing chronic diseases. Health system actors echoed these concerns, linking stockouts to broader system readiness.

Concerns also extended to the perceived quality of free government-issued medications. Some community members believed they were not as effective as those purchased privately. *“Don’t take the free medicine, because the government’s free medicine is not effective.”* A community member commented in an FGD, with others nodding in agreement. Another similarly noted, *“The health post doesn’t provide very good medicine, they give cheap medicine. If the medicine is a bit more expensive, they suggest buying it outside… They give some other, lighter version. That doesn’t work for me.”*

A few participants shared instances where doctors at government hospitals discouraged the use of government-provided medications, claiming they were ineffective. *“Even if we go there [government hospitals] and ask for government medicine, they [doctors] still tell us that the private one is better,”* a community member noted. Some also mentioned that government doctors informally directed them to seek care at those doctors’ private practices instead.

While distrust was the dominant sentiment, a few participants noted some improvements in government health services. One individual shared that the recent availability of higher-quality medications at the health post could gradually rebuild trust if consistently maintained, *“I also saw some good antibiotics there recently. When I asked, they said they just started providing it. If they consistently provide good medicines, people might start believing in the health post again.”* Others concurred that they might visit health posts more often if their preferred medicines are always available.

##### II. Financial barriers and insurance challenges

While Nepal’s government health insurance scheme provides subsidized medicines, participants frequently encountered problems. Several community members reported that the medicines covered under insurance were not the ones they needed, or that accessing insured services required additional costs and bureaucratic hurdles. A participant noted, *“They provide cheaper medicines but not the expensive ones. For example, they don’t give medicines that cost 24–25 rupees per tablet.”*

Some community members also raised concerns about transparency and mismanagement in how insured medicines were distributed. *“Cholesterol medicine wasn’t covered under the health insurance scheme. When I asked, they said it wasn’t available. But when I contacted someone I knew at Gandaki Hospital, they gave me the medicine. It costs 3500. When I checked, it was actually covered under insurance,*” a community member commented in an FGD. Another added, “*We often don’t know what’s covered under insurance. Staff keep medicines for their acquaintances.”*

For uninsured individuals, the financial burden was even greater, leading some to borrow money or take loans to afford basic prescriptions.

##### III. Patient experience

Negative interactions with healthcare providers further eroded trust. Many participants described government hospital staff as unapproachable, indifferent, or unwilling to communicate effectively with patients. One community member said, *“The thing is, if you go to a government hospital to seek consultation, they can’t even speak properly… That’s the main issue. Even when you go to the hospital, they can’t talk to the patient properly, explain things well, and provide the correct medication.”* Another echoed these frustrations, noting, *“It’s poorly managed. Some staff members don’t even let you speak. They just say, “We don’t have the medicine. Go.”*

Long wait times and bureaucratic inefficiencies create additional hurdles. As one participant recounted, “Today, for example, you have to stand in line to get the ticket, and the timing is never convenient. Government places are even less punctual. The doctor doesn’t come until 10:30 AM. Then, after getting the ticket and running around from one counter to another, you won’t finish before 3 PM”.

Perceptions of discrimination based on socioeconomic status were common, with poorer patients often feeling neglected while wealthier individuals received preferential treatment and access to better services. *“They don’t do anything there; they only discriminate between the rich and the poor,”* a participant from a rural community stated, while another described witnessing disparities in treatment, where two patients with the same illness received vastly different levels of care—one receiving all necessary medicines while the other was given only minimal support and sent outside to buy additional medications.

Some accounts were particularly severe, with community members particularly from rural populations in the peri-urban areas expressing strong distrust in certain facilities. *“The health post is there… you saw it. They try to kill people there,”* a respondent commented.

#### Private health facility preference

Due to persistent frustrations with government health services, many participants expressed a strong preference for private providers despite the financial burden.

##### I. Accessibility and convenience

Private facilities were viewed as more accessible, efficient, and patient-centered. Participants described shorter wait times, better follow-up practices, and easier appointment scheduling compared to government hospitals. As one participant explained, *“[Government] hospitals are crowded. I go to a private clinic where an army doctor does a complete check-up.”*

For those living in remote areas, physical distance compounded the challenges of accessing government health posts. Participants noted that facilities were often located far away, sometimes requiring long uphill walks or travel by vehicle, which discouraged use. An FCHV commented in a focus group, “For my community, the health post is very far. Most people don’t even know where it is!” And another added, “If it was here, I would go, but I have to climb the hill, which takes about an hour.”

Participants often expressed loyalty to private doctors they had been consulting for years, preferring continuity of care over switching to government hospitals. A community member shared, *“Yes, I’ve had severe headaches since before, so I didn’t stop seeing that doctor.”*

However, a few health workers highlighted that government health posts remained a primary option for some residents. One FCHV noted that their health post was centrally located and saw a steady daily patient load of around 40–45 people. Additionally, an FB-CHW mentioned that in areas with fewer private clinics, people tended to rely on health posts for their healthcare needs.

##### II. Financial burden of private healthcare

While many community members felt that private facilities had better service, the costs could be steep. Some participants discussed how people, even those with limited resources, would go to great lengths to afford private healthcare: *“Even those who can’t afford it [private health facilities] also go due to lack of any choice… They might even sell a buffalo to pay.”*

Some community members and FCHVs noted that individuals from lower-income groups often had to rely on government health services due to financial constraints. For these populations, health posts were viewed as the primary, and sometimes the only, accessible option for medical care.

#### Workload and motivation: FB-CHWs

While community members expressed concerns about quality and tended to blame facility staff, facility based CHWs (FB-CHWs) pointed to systemic concerns about their increasing workload, limited staffing, and the challenges they face in balancing multiple responsibilities.

FB-CHWs described their work as diverse and demanding, encompassing both clinical services and community outreach. Many expressed that their responsibilities extend far beyond basic patient care. Their daily duties include patient check-ups, medication refilling and health counseling, but they also run additional programs such as vaccination clinics, outreach services, family planning, tuberculosis and malaria control, and school health initiatives. These tasks often require traveling to remote areas and ensuring that underserved populations receive essential care. Additionally, they are responsible for compiling service data and ensuring accurate reporting.

One of the biggest challenges reported by FB-CHWs was the shortage of health workers, which increased their workload and created additional stress. Many health posts operated with only three or four staff members, forcing FB-CHWs to manage multiple services simultaneously. An FB-CHW described the strain of working with a limited team, *“We are two [at the health post]… We do feel work overload. There is insufficient staff… We both have to look after all these services. One has to manage OPD services, another has to manage family planning and antenatal checkups. We also have to manage the administrative part. So, there is massive workload.”*

When a staff member was absent, the burden intensified, “*Sometimes, when we are alone, we face some problems. If patient-flow is high sometimes, if both of us are there, then we take our patients and do our tasks separately. But it is quite difficult if only one of us is present at the health post.”*

Many FB-CHWs felt the strain of balancing multiple roles with limited staffing. Some noted that their workload has increased as NCD cases rise in the community, requiring them to incorporate hypertension and diabetes screenings into their already full schedules. One FB-CHW shared, *“Previously, we only had a blood pressure measuring device. We used to check blood pressure only for those showing symptoms. But now, we measure the blood pressure of every patient who visits us because NCDs have been increasing significantly.”*

While some respondents believed the workload at health posts was manageable, many FB-CHWs emphasized that without additional staff or time, participating in new NCD interventions would be difficult. Despite these challenges, FB-CHWs remain committed to their work. However, timely salary payments and increased staffing were highlighted as critical factors in maintaining motivation.

### 4. FCHVs in NCD management

#### General perceptions of FCHVs

Community members generally recognized FCHVs as familiar figures in their neighborhoods, particularly for their work in maternal and child health. Their visibility in vaccination campaigns and health education initiatives contributed to a degree of trust. However, many participants noted that FCHVs primarily focused on child health, such as polio vaccinations, with limited involvement in addressing adult healthcare needs.

While FCHVs’ contributions were generally valued, perceptions varied regarding the depth and consistency of their engagement, with some viewing their role as symbolic, and visits as sporadic and dependent on specific government programs. *“They do visit during festivals like Dashain and Tihar when there’s a need to give polio drops. Otherwise, we don’t see them*,” a community member noted. A few individuals expressed skepticism about their contributions, suggesting that their work was minimal or that they did not provide substantial services.

Under Nepal’s national health system, FCHVs are formally responsible for delivering maternal, newborn, and child health services. NCD-related tasks, such as screening and counselling for hypertension or diabetes, are not part of their standard government assigned responsibilities and are generally introduced through time-limited programs led by non-governmental organizations, academic institutions, or international development partners. Consequently, community exposure to FCHVs’ role in NCD care has been shaped by whether such external programs were active in their area.

#### FCHVs’ emerging role in NCDs

While FCHVs reported an increasing level of trust and engagement in their NCD-related activities, community perceptions ranged from appreciation to skepticism about their capabilities.

##### I. FCHVs’ perspectives

FCHVs described how initial community skepticism about their ability to perform NCD screenings had gradually shifted toward trust and recognition. *“In the past, people didn’t value health checks like blood pressure and sugar tests. They’d mock us, asking, ‘What do you measure with your gadgets?’ But now, they seek us out,”* one FCHV explained.

Trust was reinforced when community members noticed that the same digital devices used by FCHVs were also being used in hospitals and health centers, and when FCHV-led screening led to confirmed diagnoses at hospitals. *“Many people would claim they didn’t have sugar [diabetes] or high blood pressure,”* one FCHV commenting, adding, *“But when we tested them and referred them to health institutions, they found out they actually had those conditions. That’s when they started believing us…*

*They came back and thanked us for detecting their issue.”* They also noted that their constant presence in the community made them well-positioned to provide early detection and referrals.

Beyond detection, FCHVs also saw their role as extending into patient follow-up. *“We volunteers also keep track. If someone is diagnosed with high blood pressure, we ask if they’re taking their medication. If they are, we advise them to consult a doctor if their condition doesn’t improve. If they’re not on medication, we refer them to a hospital or health post for further advice,”* one noted.

However, despite their growing role in NCD management, some FCHVs expressed frustration that their work was not always valued. A few noted that community members sometimes resisted their health assessments, questioning the purpose of screening without immediate treatment. *“Some people said things like, ‘If you’re not going to give me medicine, why are you even testing me?’ Now, some people are asking when I will start again, urging me to resume checking.”* an FCHV commented. Others shared that having access to their own glucometers and blood pressure monitors would strengthen their ability to provide effective services.

##### II. Community perceptions of FCHVs in NCD care

Community perceptions of FCHVs’ role in NCD care were mixed, including both optimism and concerns. Some participants appreciated FCHVs’ expanding role and suggested that more frequent home visits for screening and counseling would be beneficial. *“For example, they could check blood pressure and tell us if medicine is available. If we knew when sugar tests were conducted, that would be helpful.”*

At the same time, skepticism persisted, particularly regarding the perceived medical expertise of FCHVs. Some questioned their ability to conduct accurate screenings for chronic conditions, suggesting that such tasks required trained medical professionals. *“They don’t visit like your [research] team does. And even if they came, they don’t know how to check blood pressure properly*,” a community member commented, and another added, *“Volunteers can’t help; they spend their days working in cow sheds, and then they come and say, ‘Let’s measure your blood pressure.’ What’s the use?”*

Concerns were also raised about the lack of clarity in communication following screenings, with some participants emphasizing the need for FCHVs to not only measure but also explain results and next steps.

As one community member stressed, *“They need to detect the disease, check the sugar, check the blood pressure… If medication is required, if the disease needs to be explained—everything. Just tying a watch and taking it off is not enough.”*

Despite some skepticism, there was agreement that with adequate training and support, FCHVs could play an important role in improving early detection and management of hypertension and diabetes at the community level. Participants, particularly from rural communities, emphasized that FCHVs’ close ties to the community positioned them well to bridge gaps in care by conducting regular screenings, ensuring follow-up, and linking individuals to health services.

##### III. Healthcare system perspectives

Facility-based CHWs (FB-CHWs) viewed FCHVs as valuable allies for expanding community-based NCD care. Their accessibility and community trust were cited as major strengths. *“FCHVs are available 24/7. But we only operate between 10 AM to 5 PM, so in terms of timing, they are always accessible, morning or evening,”* a FB-CHW noted.

FB-CHWs particularly relied on FCHVs for community mobilization and outreach, especially during national health campaigns. *“In earlier days, during door-to-door programs, people wouldn’t show up when called. FCHVs know every household personally, so they manage to call them somehow. When we go alone, people don’t recognize us,”* an FB-CHW noted.

Despite their mobilization strengths, FB-CHWs said they thought that FCHVs’ role should remain primarily informational, rather than clinical. *“Their [FCHVs] job is to inform the community when a program is coming, to let people know. But when it comes to actually conducting the [health] program, that’s our job. They don’t have that capacity. Our volunteers have a little education,”* one AHW explained.

Still, FB-CHWs acknowledged that FCHVs had gained valuable skills through formal training programs. *“Initially, they weren’t aware of these [NCD] conditions. Later, after receiving training from Nepal Vikas Samaj, they gained knowledge about these diseases. They started measuring glucose levels with glucometers and checking blood pressure. If the readings were high, they would refer the patients to the health post,”* a FB-CHW explained.

However, they also identified the need for more structured support systems, sustained training and stronger integration with the healthcare system to maximize FCHVs’ potential impact. *“They would benefit if they had their own tools to measure blood sugar and blood pressure. If they were trained and equipped to do so, they could measure these for people nearby,”* an FB-CHW commented. Others noted that while FCHVs were trusted within their communities, their role in NCD care was limited by gaps in medical knowledge. They also questioned the sustainability of relying on FCHVs for NCD care, as one participant noted, *“Looking at sustainability, FCHVs might not last long because most of them are not highly educated. In the future, we might not get anyone ready to be FCHVs, as people may not want to volunteer anymore.”*

Stakeholders across government and academic institutions emphasized the value of FCHVs, emphasizing the centrality and deep integration of FCHVs in community-level health service delivery. A public health officer also highlighted FCHVs’ evolving scope of work, noting their increasing involvement in NCD prevention and management through health education and basic screening. *“People really look at FCHVs positively… they have worked in maternal health, so now people look at them with respect,”* an organization representative emphasized their deep ties to the community and natural ability to engage in NCD outreach.

While most stakeholders expressed high regard for FCHVs, some also noted contextual limitations. One participant explained that trust in FCHVs was stronger in rural areas compared to urban ones, where wealthier and more educated households might be less receptive to volunteer-led care. A public health officer noted education and the aging of the FCHV workforce without corresponding recruitment of new volunteers as a critical constraint*, “Older volunteers have retired… and the center is not recruiting new ones.”* He proposed potential policy solutions to address growing burden of NCDs, including the recruitment of replacement volunteers, deploying Auxiliary Nurse Midwives (a type of FB-CHW) at the community level and expanding the model to include male community health volunteers.

Overall, while there was broad support for involving FCHVs in NCD management, several structural and systemic challenges were seen as critical to address before scaling such approaches. The following section unpacks these implementation barriers, including workforce constraints, training, and compensation and sustainability.

### 5. Health system constraints to community-based NCD care

#### Compensation

Compensation for FCHVs remains a key issue, with volunteers frequently expressing concerns over low and inconsistent allowances. While many remained committed to serving their communities, they emphasized that the current system often created financial strain and did not adequately reflect the time and effort required for community health work.

FCHVs nearly universally commented that remuneration did not fairly compensate for the work they do. FCHVs reported that attending trainings and health activities often required them to forego agricultural work or other paid employment, exacerbating financial hardship. They pointed out that they had to spend money on transportation costs, and that the opportunity costs of FCHV work were severe.

These constraints had implications for patient care. For example, a lack of financial support for mobile communication expenses was a barrier to effective patient follow-up: *“If we need to make calls, perhaps a provision for a SIM card or phone expenses could be made. If we are working voluntarily, it’s difficult to spend from our own pockets,”* one FCHV commented.

Stakeholders emphasized the need for structured financial incentives to maintain FCHV motivation. A hospital superintendent added*, “They are not satisfied. Their daily complaint is that they are not satisfied with what they got… The motivation to engage them is financial. That’s what’s needed now.”*

A national-level stakeholder emphasized the broader policy dilemma, noting the importance of offering motivational incentives even within the voluntary framework. Similarly, another stressed the importance of both financial and non-financial recognition as a strategy for retaining FCHV engagement*, “We can’t give them salary… but we must motivate them through some form of incentives. Free insurance, free medicine, or even lunch and transport costs during program days — these things should be offered.”*

#### Training and resource limitations

Most FCHVs appeared willing to take on additional tasks related to NCD management. However, health care workers, both FCHVs and FB-CHWs, noted that while they have received some training for NCD management, it is often infrequent and insufficient to equip them with the necessary knowledge and skills to manage NCDs effectively.

A few participants also highlighted that past training initiatives, while effective, were limited in scope, often targeting a select group of individuals rather than being made widely available. In some cases, training programs were discontinued after the initial rollout. Others emphasized the need for refresher training, as well as ongoing supervision and support to help FCHVs effectively apply their training in practice.

Several also emphasized that sustaining training and supplies required systemic integration of task-sharing with FCVHs.As one policymaker noted, *“The most important aspect of sustainability is integrating it [task-sharing intervention] into the system.”* He added, *“The main thing is to use the evidence to convince policymakers since they are the ones who ultimately implement it. Health involves various partners, and we need to demonstrate to them—especially in the health system—that this is effective.”* Additionally, many FB-CHWs struggled with stock-outs of essential medical supplies, particularly glucometers, test strips, and medications, and FCHVs reported that they lacked the necessary tools— such as functioning BP monitors and glucometers—to effectively implement what they learned in training. They emphasized ensuring continuous provision of materials and resources to allow uninterrupted service provision to the community.

The stock-outs could often last up to three or four months. One FB-CHW explained: *“Since we don’t have the means to purchase the kits ourselves, we rely on the government or other organizations to supply them. If they provide the kits, we can offer the service. For instance, there was a time when we experienced a stock-out. Even if we have the machine, we can’t work without the kits.”*

Another FB-CHW described the ongoing challenges of inconsistent supply chains*, “What was available before is no longer available now. When it’s out of stock, do patients still come in? Yes. Do they scold us? Yes. They say, ‘You didn’t ask for it [medication supply].’ But we did.”*

Despite these challenges, some facilities reported better stock management, indicating variability in supply distribution. One FB-CHW noted*, “No. It’s never gotten out of stock till date. Not even for one day.”* FCHVs also reported facing logistical challenges due to outdated or poorly designed equipment. *“Can we have better-designed bags for carrying our materials? Something that can be carried easily wherever we go. A backpack style would be ideal.”* an FCHV requested during a discussion on home visits.

#### Sustainable resource management

Ensuring the sustainability of community-based NCD interventions emerged as a major concern among both health workers and stakeholders. Participants emphasized that short-term external support, while helpful, would not address the deeper structural issues that limit consistent service delivery.

The concerns about resource continuity were common were often linked to concerns about sustainability. One FCHV commented, *“Sometimes, we even question whether we should continue with this work. How can we come closer to making this system work? If you provide training, ensure availability of common medicines for sugar and blood pressure patients at these posts. If such systems were in place, things would improve a lot.”*

In the absence of reliable government support, partnerships with external partners—such as NGOs and provincial health offices—were seen as a potential solution for filling supply gaps. *“If we can try to link up with some other projects from other countries or provincial governments, they can also supply additional drugs like metformin, Amlod, and other essential medicines.”* a FB-CHW suggested.

A government officer emphasized the importance of systemic reforms to improve access to medicines and expand insurance coverage for patients. Referring to common complaints raised by patients, particularly those whose prescriptions involved multi-composition drugs not currently covered under insurance resulting in high out-of-pocket expenditures, he proposed multiple strategies to reduce this burden. He noted, *“If we can supply composition-based medicines, or at the metropolitan level establish affordable pharmacies – maybe two or three outlets that offer medicines 20–30% cheaper than other places – or at the national level, expand the scope of insurance, then we can make some progress. Expanding the insurance limit from NPR 100,000 to NPR 500,000 and adding those multiple-composition medicines to the current list could make a significant difference. Right now, there are about 1,100 items on the list; if we can increase that to 1,500–1,700, those cases could be covered.”* He also cautioned that as NCD burden and treatment complexity grow, addressing spending patterns and patient affordability will become an even more pressing challenge over the next five to seven years.

Participants also stressed that achieving sustainability requires more than supplies and insurance reform. Government ownership at the municipal and ward levels was viewed as critical. A stakeholder emphasized that elected officials needed to recognize the long-term value of community-based NCD interventions delivered through FCHVs in order to prioritize them in local budgets and planning. *“At the municipal level, not just the staff but also the mayors or municipal chiefs, or health coordinators from the municipalities, should recognize its effectiveness… They should think, ‘For this, we need to allocate a budget and communicate with the female health volunteers.’”*

Without active government engagement and integration into existing budgets, participants cautioned that successful interventions would likely not be sustained once external projects ended. *“If the program is still running two to three years after you leave, it shows that the program you initiated was effective… Efforts and programs should be implemented to make that accountable government take even more ownership.”*

Given the resource and staffing challenges in community-based care, participants shared perspectives on the potential of digital tools to supplement existing efforts. The final section examines perceptions of mHealth strategies—including SMS and audio messages—and their feasibility in supporting NCD management.

### 6. Feasibility of mHealth strategies

#### General perception

Community members and healthcare workers had mixed perceptions about technology-based interventions, such as SMS and phone call reminders, for NCD management. While many participants saw the potential benefits of such outreach strategies, others expressed concerns about accessibility, effectiveness, and user behavior.

##### I. Perceived benefits

Many participants viewed virtual reminders as a convenient and helpful tool for promoting regular health check-ups and medication adherence. FCHVs noted that reminders could help individuals remember their appointments and encourage them to seek care on time.

Community members living in peri-urban areas expressed enthusiasm for receiving automated health reminders, comparing them to existing festive greeting messages they receive during festivals. They suggested that such messages could serve as effective prompts for seeking healthcare. One noted, *“If you send volunteers door-to-door, some houses might not have anyone at home, or they might miss someone. But a mobile phone is almost always present with at least one person in the household. When a message arrives, they look at it once, press it, and even those who can’t read will put it to their ear and listen. They will understand the message and receive the information.”*

Other community members expressed more cautious support, suggesting that phone call reminders would be even more effective than text messages. *“If I get a call saying, ‘It’s time to check your blood pressure,’ that would be helpful. Then you can plan and make time.”* They also emphasized that in-person visits from the FCHV remained an important and welcomed form of engagement.

FB-CHWs also supported the idea of mobile messaging or calling. One commented, *“That would be great. Everyone in the household would have the chance to know [health information], including those who can’t visit the health post. Nowadays, almost everyone has a mobile phone.”*

Several stakeholders also expressed strong support for mHealth interventions like SMS and audio reminders. A senior consultant from the Ministry of Health and Population’s Epidemiology and Disease Control Division (EDCD) cited the successful use of mobile alerts in maternal health campaigns and emphasized that a hybrid approach—combining digital messages with in-person visits—would be most effective in sustaining community engagement. An international organization representative echoed this support, noting that messages personalized with FCHV names and sent from trusted health institutions could enhance credibility.

##### II. Challenges and limitations of SMS-based interventions

Despite the potential benefits, key concerns included literacy and phone accessibility, feasibility and long-term sustainability. *“Not everyone has a phone, and in some households, only one person might have one… In such cases, the message might not reach the intended person unless someone tells them.”* an FCHV explained in an FGD. Another added, *“Some people don’t know how to read.”* A FB-CHW further emphasized*, “Some [community members] are very poor. They can’t afford repairing their phones once they get damaged.”*

Healthcare workers also raised concerns that elderly individuals, who represent a significant portion of the NCD patient population, may not engage with SMS interventions. *“In villages, it’s mostly elderly people who stay behind. Their sons and daughters, who could understand such things, don’t live with them. So, I don’t think text messages will help them,”* a FB-CHW noted.

Even among those with phones, several community members noted that busy daily routines meant messages might be overlooked. *“For those of us who might not understand messages, having someone come in person is always better,”* a community member observed.

There was also the question of who would have time to send the messages. Many health workers expressed that while they recognized the value of phone-based reminders for NCD management, staffing shortages and high workloads made it difficult for them to implement such interventions directly. *“Doing it personally will be difficult for us,”* one FB-CHW explained, “*We already have so much work pressure here, we don’t have enough time for phone calls.”* A differing perspective emerged from another FB-CHW, who believed the workload was manageable, and FB-CHWs should be able to take on additional responsibilities: *“I think they would be able to do it because there’s not much workload at health posts, and it’s not very crowded. In areas like Lekhnath, there are about 30–40 patients a day, which is not too many.”* This viewpoint contrasts with the dominant narrative of overworked staff and insufficient personnel, highlighting potential variability in workload across different health facilities.

Stakeholders from the government and telecom sectors raised feasibility concerns related to SMS-only approaches. A representative from Nepal Telecom noted that while SMS can be a useful tool, its reach is limited in rural areas due to literacy barriers. He also emphasized that technical and operational challenges must be addressed to ensure long-term sustainability.

One major constraint is the need for an automated system capable of managing message delivery logistics over time. Manual approaches may work temporarily but are not scalable or sustainable for long-term health programs, *“You’ll need to manage who gets the message, when, and how often it’s sent,”* he explained.

He further described the additional requirements of using recorded voice calls, *“If you want to send an audio file, you need data. The person must have their data turned on. Or, they must have an internet connection for that.”*

Beyond audience-level limitations, a robust backend platform that can maintain user profiles, communication preferences, and track delivery logs are required. *“If you want to go for the long term, you need to develop a system… store information about all the people in Pokhara, their mobile numbers, and details about their conditions… and through which medium it would be most appropriate.”* he explained. Finally, regulatory and legal constraints present additional challenges for long-term intervention delivery. While Nepal Telecom offers bulk SMS and Interactive Voice Response (IVR) services, regulatory guidelines require individual user consent for direct communication from service providers. Privacy issues, particularly related to storing personal health information and mobile numbers, must be carefully addressed when designing sustainable SMS interventions.

##### III. Preference for in-person communication

Some participants felt that direct, in-person communication from FCHVs or healthcare providers would be more effective than text messages. They emphasized that face-to-face interactions allow for clarification, personalized advice, and greater accountability.

FCHVs and FB-CHWs also noted that while mobile communication has made follow-ups easier, in-person visits remain crucial for ensuring that health messages are understood and acted upon. They explained that face-to-face interactions often led to better engagement and compliance. One FCHV shared, *“When I send a message to a pregnant woman, it says something like, ‘It’s been this many months, you need to go for a checkup.’ But when I visit her in person, she actually goes.”*

##### IV. Alternative communication strategies

Many participants expressed a strong preference for phone calls over text messages. They felt that a direct phone call would be more engaging and easier to understand, especially for elderly individuals or those with limited literacy.

Health workers also emphasized that phone calls create a sense of care and responsibility, making patients feel valued and more likely to follow through with health recommendations. *“When we refer a complicated case from our hospital and then follow up with a call to ask about their condition, the patients respond very positively and are happy to hear from us. They feel like we cared enough to call them, which creates a very positive impression.”* a FB-CHW commented.

Audio or recorded voice messages were viewed as an effective alternative to text messages, where participants could listen to the message and gain the necessary information. *“Audio has been more effective than reading. Even those who cannot read can listen to it on a mobile device and get the information they need,”* a community member noted in an FGD, and others agreed.

Several community members also suggested that recorded messages from trained professionals, such as doctors or nurses, would enhance credibility and encourage adherence. One commented, *“Providing information through female health volunteers is good, but wouldn’t it be better if the audio recordings were made with experts like doctors or nurses?”*

Some also proposed using WhatsApp or other social media platforms to disseminate health information, allowing for group discussions and engagement. Healthcare workers recommended that technology-based interventions be supplemented with visual aids, such as flip charts and videos, to improve comprehension and enhance engagement.

While mHealth strategies were generally perceived as helpful for reminders and follow-up, participants emphasized that their effectiveness would depend on access to mobile phones, digital literacy, and trust in phone-based communication. Importantly, health workers and FCHVs noted that mHealth interventions alone could not address the broader challenges faced in NCD management. Persistent issues — including staffing shortages, inconsistent availability of medicines and equipment, and limited training opportunities — were seen as critical barriers that would need to be addressed alongside digital strategies to ensure the feasibility and sustainability of community-based interventions like SCALE-NCD.

## Discussion

While community members demonstrated awareness of hypertension, diabetes, and smoking as key health concerns, multiple factors shaped their risk exposure and health-seeking behaviors. Once diagnosed, most participants reported consistent adherence to prescribed medications; however, challenges in sustaining lifestyle modifications and treatment adherence included financial constraints, concerns about long-term medication use, and limited access to services. Trust in government health services emerged, not surprisingly, as a central determinant of where and how people seek NCD care (17,18). Stock-outs of essential medicines, perceived poor quality of free medications, and negative provider interactions contribute to a strong preference for private healthcare despite its financial burden. Institutional trust—built through reliable, respectful, and competent system performance—is essential to increasing health care service utilization. Our findings emphasize the need to strengthen government health services in parallel with community-based efforts to prevent fragmentation and support long-term engagement in care.

Against this backdrop, FCHVs were recognized as trusted, accessible figures within their communities particularly for maternal and child health, and increasingly, for their role in NCD screening. Initial skepticism towards FCHVs’ ability to conduct blood pressure and glucose monitoring appeared to diminish over time, especially when their screening results were confirmed by FB-CHWs. Importantly, community acceptance of FCHVs in NCD management was closely tied to perceptions of their training, skill level, and ability to explain results and provide appropriate follow-up guidance. Reliable provision of medical equipment and supplies, consistent availability of medications, and robust training programs were seen as critical enablers for both FCHVs and FB-CHWs. This echoes other studies emphasizing the need for training and support for FCHVs (11,18,19). Ensuring that FCHVs are equipped not only with tools and a stable supply chain, but also with communication skills and technical capacity will be essential to the success of task shifting for NCDs.

While participants in our study strongly supported the expanding role of FCHVs in NCD prevention and screening, concerns around workload, motivation, and sustainability of a voluntary model were raised. Existing research on the FCHV program has highlighted the growing misalignment between expectations of FCHV performance and the limited incentives provided to support their work (20,21). Recent literature suggests that expectations for volunteers to deliver increasingly time-bound and medically technical services — such as blood pressure screening — without corresponding compensation can erode both morale and legitimacy. While many FCHVs remain motivated by community service, religious merit, and social recognition, stakeholders have noted that expanding responsibilities — including disaster response and NCD screening — may require rethinking incentive structures. Importantly, incentives should be sustainable and transparent, and aligned with both FCHVs’ motivations and evolving health system demands.

The ideal solution is to offer a salary, but even in the absence of a salary, supports such as advanced training, consistent and supportive supervision, reliable availability of supplies, activity-based allowances, retirement stipends, and priority access to government services can be important supports and motivators for CHWs (22–26). Visible incentives such as transport reimbursement, free healthcare, representation in local health management committees, and public acknowledgment have been noted as ways to support motivation while preserving the community-based ethos of the program (20,27). Efforts to sustain the FCHV program must consider not only equitable and consistent incentive structures, but also improved supervision, workload boundaries, and respect from the broader health system (28). As task-sharing for NCDs continues to expand, aligning program demands with FCHVs’ motivations, expectations, and lived experiences will be critical to long-term effectiveness.

The use of mHealth strategies such as SMS and phone call reminders emerged as a promising but complex intervention component. Digital communication was widely seen as helpful for reminders and behavior change support, but concerns around literacy, mobile phone penetration, and message comprehension — especially among elderly, low-income, and peri-urban populations — raised questions of digital equity. Stakeholders highlighted the need for audio-based formats, simplified messages, and multimodal delivery mechanisms. These concerns align with prior evidence from mHealth interventions in LMICs, which emphasize the importance of user-centered design, technology literacy, and inclusive access strategies for scaling digital health solutions (29,30). Without such adaptations, digital health solutions risk limited uptake or benefit among the populations most in need of support.

Furthermore, sustainability of such strategies hinges not only on end-user uptake but also on backend infrastructure and institutional coordination. Our findings suggest that message delivery systems require significant setup, automation, and regulatory compliance, including user consent, approval from telecom authorities, and integration with public health tracking systems. Successful integration of mHealth into community-based NCD care will depend on addressing both technological and human-level barriers — ensuring that digital strategies complement, rather than replace, the interpersonal trust and outreach provided by FCHVs.

Finally, a key theme that emerged in this study is that long-term sustainability hinges on systemic integration and government ownership. While donor-funded programs can catalyze innovation, they often risk collapse unless institutionalized within local systems. To ensure continuity, stakeholders such as municipal mayors, ward chairs, and health coordinators should be engaged early—not just as implementers, but as long-term stewards (21).

These findings have several implications for task-sharing and mHealth as strategies for NCD care in Nepal. Special attention is needed to reach populations at risk of being left behind — such as lower-income individuals and those in peri-urban or remote areas — who face financial and geographic barriers to care. Strengthening government health services, improving medicine availability, and investing in patient-centered care are critical complements community-based efforts and help rebuild trust in public health systems. Addressing workforce shortages, ensuring equipment availability, and developing supportive supervision structures will be essential to sustain task-sharing models. And importantly, building a supportive environment for FCHVs—through tools, training, recognition, and community legitimacy—remains foundational to intervention success.

As Nepal confronts a growing burden of NCDs, integrating community health workers like FCHVs into NCD prevention and management presents a promising strategy to expand reach and improve health equity. However, success will depend not only on building FCHVs’ technical capacity but also on addressing systemic barriers within the health system, ensuring sustained support, and fostering trust among communities.

## Data Availability

Full interview and focus group transcripts cannot be shared publicly to protect participant confidentiality. Relevant de-identified excerpts are included within the paper. Additional anonymized data supporting the findings of this study are available from the corresponding author (DN) upon reasonable request.

